# Sexually transmitted infection risks and symptoms among women who started selling sex before the age of 18 in five cities in Cameroon

**DOI:** 10.1101/2022.05.27.22275267

**Authors:** Ashley Grosso, Anna L. Bowring, Iliassou Mfochive Njindam, Michele R. Decker, Carrie Lyons, Amrita Rao, Ubald Tamoufe, Guy H. Fako, Ghislaine Fouda, Daniel Levitt, Gnilane Turpin, Serge C. Billong, Anne Cécile Zoung-Kanyi Bissek, Oudou Njoya, Stefan Baral

**Affiliations:** Institute for Health, Health Care Policy and Aging Research, Rutgers University, 112 Paterson Street New Brunswick, NJ 08901 United States; Department of Epidemiology, Johns Hopkins University, 615 N Wolfe St Baltimore, MD 21205 United States; Burnet Institute, 85 Commercial Road, Melbourne VIC, 3004 Australia; Department of Population, Family, and Reproductive Health, Johns Hopkins University, 615 N Wolfe St Baltimore, MD 21205 United States; Metabiota, Avenue Mvog-Fouda Ada, Av 1.085, Carrefour Intendance BP 15939 Yaoundé, Cameroon; CARE Cameroon, Villa La Rose (3è étage, Av. Churchill, Yaoundé, Cameroon; CARE USA, 115 Broadway, 5th floor, New York, NY 10006 United States; Groupe Technique Central, CNLS, Rue Henri Dunant, Yaoundé, Cameroon; Department of Operational Research, Ministry of Health, Road 3038, Quartier du Lac, Yaoundé, Cameroon; Department of Internal Medicine, Yaoundé University Hospital Center, Joseph Tchooungui Akoa, Yaoundé, Cameroon

**Keywords:** sex workers, Cameroon, sexually transmitted diseases, HIV testing, adolescent

## Abstract

**Purpose:** Many adolescents under 18 years old who sell sex are at elevated risk for sexually transmitted infection (STI) acquisition, which may persist into adulthood. There has been limited study of the burden of the risks and vulnerabilities among women who started selling sex as adolescents across Sub-Saharan Africa.

**Methods:** Adult female sex workers (FSW) recruited through respondent-driven sampling in five cities in Cameroon from December 2015 to October 2016 completed a questionnaire and human immunodeficiency virus (HIV) and syphilis testing. Multivariable logistic regression analysis controlling for age was used to identify factors associated with reporting selling sex before age 18.

**Results:** Selling sex before age 18 was reported by 11.5% (256/2,220) of FSW. Initiation of selling sex as an adolescent was positively associated with experiencing dysuria (adjusted odds ratio [aOR]:1.50, 95% confidence interval [CI]:1.08-2.10) or genital warts (aOR:1.78, 95% CI:1.08-2.94) and negatively associated with prior recent testing for HIV (aOR:0.71, 95% CI:0.53-0.96) or STIs (aOR:0.65, 95% CI:0.44-0.96). Consistent condom use with clients was negatively associated with early initiation of selling sex (aOR:0.58, 95% CI:0.42-0.80), while experience of recent sexual violence was positively associated with early initiation (aOR:1.74, 95% CI:1.15-2.63). There were no independent significant differences in HIV (24.5%) or syphilis (8.4%) prevalence.

**Conclusions:** Given the limited use of HIV and STI testing services by women who sold sex as adolescents, the prevalence of forced sex, condomless sex, and STI symptoms were high. Programs serving FSW should more vigorously aim to serve adolescents and adults who began selling sex early.

## Main text

It has been posited that adolescent girls and young women in some settings are neglected key populations at particularly high risk of HIV infection.(Dellar, Dlamini, & Karim, 2015) Vulnerability indices have been developed to assess HIV risks among adolescent girls and young women to guide implementation of large-scale programs including the Determined Resilient Empowered AIDS-free Mentored and Safe (DREAMS) initiative.(Han, 2017) Through this research and other studies, transactional sex has been identified as a factor potentially leading to elevated HIV risk for this population.(Chiyaka et al., 2018; Michele R. Decker et al., 2014; Head, Eaton, Lloyd, McLaughlin, & Davies-Cole, 2021; Martin et al., 2021; Stoebenau, Kyegombe, Bingenheimer, Ddumba-Nyanzi, & Mulindwa, 2019) International guidance does not differentiate transactional sex from sex work, and these definitions are often contentious. Terminology is also controversial regarding adolescents under 18 years old who sell sex for money, many of whom are at elevated risk for sexually transmitted infection (STI) acquisition. This risk is particularly concerning in sub-Saharan Africa, where many settings have high HIV and STI prevalence among reproductive-aged populations.(Newman et al., 2015) For example, in Mozambique, compared to adult female sex workers (FSW), 15- to 17-year-olds selling sex had a higher prevalence of recent STI symptoms.(Inguane et al., 2015) STIs are associated with increased vulnerability to HIV, pelvic inflammatory disease, and infertility.(Mwatelah, McKinnon, Baxter, Abdool Karim, & Abdool Karim, 2019) This higher prevalence of STIs including HIV can persist into adulthood. Among adult FSW in Cote d’Ivoire, initiation of selling sex before age 18 rather than as an adult was associated with testing positive for HIV.(A. L. Grosso, C; Diouf, D; Liestman, B; Ezouatchi, R; Thiam-Niangoin, M; Abo, K; Drame, FM; Bamba, A; Tety, J; Obodou, E; Ba, I; and Baral, S., 2017)

Biological and behavioral factors likely contribute to elevated risk among youth who sell sex. Biologically, adolescent girls who sell sex are vulnerable to STIs including HIV due to larger areas of cervical ectopy and trauma to an immature genital tract increasing the likelihood of tearing during sex.(Silverman, 2011) Behaviorally, part of this increased risk is likely due to condomless sex. Adolescent girls who sell sex in Mozambique more commonly reported having condomless sex with their last client and less commonly reported recently receiving free condoms compared to adult FSW.(Inguane et al., 2015) These barriers to condom use experienced as youth may continue in adulthood. Among adult FSW, initiation of selling sex during adolescence rather than adulthood has been shown to be positively associated with lower condom negotiation self-efficacy in Kenya,(Parcesepe, L’engle, et al., 2016) reporting clients removed condoms or offered more money for condomless sex in Burkina Faso,(A. L. Grosso et al., 2015) and receiving fewer free condoms and avoiding carrying condoms out of fear of trouble with police in Lesotho.(A. B. Grosso, Shianne; Mothopeng, Tampose; Sweitzer, Stephanie; Nkonyana, John; Mpooa, Nkomile; Taruberekera, Noah; and Baral, Stefan., 2018)

Experience of physical and sexual violence due to gender-based and age-based power differentials may also contribute to the higher STI risk among adolescent girls who sell sex.(Silverman, 2011) Over half of children aged 11-17 years old who sold sex in Benin, Burkina Faso, and Niger reported they had experienced violence from clients.(Hounmenou, 2017a) Early abuse could shape future exposure to violence among FSW. FSW in Kenya who started selling sex as adolescents had higher odds than those who started selling sex as adults of experiencing recent violence from a client, endorsing violence-related gender norms, and having less violence-related self-efficacy.(Parcesepe, L’Engle, et al., 2016) In Lesotho, experiencing recent physical violence was more common among FSW who started selling sex as adolescents than among those who started as adults.(A. B. Grosso, Shianne; Mothopeng, Tampose; Sweitzer, Stephanie; Nkonyana, John; Mpooa, Nkomile; Taruberekera, Noah; and Baral, Stefan., 2018)

Despite greater vulnerability to HIV and other STIs, both adolescents who sell sex(Inguane et al., 2015) and adults who started selling sex as adolescents(A. L. Grosso et al., 2015) report less frequent testing than those who sell sex only as adults. In contrast, however, out-of-school adolescent girls and young women involved in transactional sex work were more likely to have tested for HIV than those not involved in transactional sex.(Han, 2017)

In Cameroon, limited research has examined STI risks among adolescents who sell sex.(Mbassa Menick, Dassa, Kenmogne, & Abanda Ngon, 2009) Pooled HIV prevalence among adult FSW across six studies in Cameroon was estimated at 23.6%.(Papworth et al., 2013) Antiretroviral treatment coverage was estimated to be only about 11% in 2013.(Holland et al., 2015) In another study, gonorrhea and chlamydia prevalence among FSW in Cameroon ranged from 10 to 13%.(Roddy et al., 1998) Other prior studies found 35.5%(Mosoko, Macauley, Zoungkanyi, Bella, & Koulla-Shiro, 2009) and 21%(Ryan, Roddy, Zekeng, Weir, & Tamoufé, 1998) of FSW in Cameroon self-reported ever having an STI. In addition, violence against FSW in Cameroon has been widely reported and associated with condomless sex.(Michele R Decker et al., 2016) Given the prevalence of STIs among FSW in Cameroon and the dearth of data on STIs among adolescents who sell sex in Cameroon, this study’s purpose was to compare prevalence of STI symptoms and risk factors among FSW who started selling sex as adolescents to FSW who started selling sex as adults.

## Methods

### Sample

Study data are from FSW recruited through respondent-driven sampling (RDS) from five cities in Cameroon from December 2015 to October 2016. At each study site one or two “seeds” (initial participants) selected through convenience sampling participated in the study and were given RDS coupons to recruit up to three peers. This process was repeated until the sample size was met. Eligible participants had been assigned female sex at birth, were 18 years old or older, reported sex work as their primary income source in the past 12 months, were capable of providing verbal informed consent, spoke and understood English or French, did not participate in a similar study in the past six months, presented an unexpired unduplicated RDS coupon (except seeds), and resided in a study city for the past three months. Participants received 2,000 FCFA (∼4 USD) as reimbursement for transportation costs and an additional 1,000 FCFA (∼2 USD) for each eligible participant they recruited who completed the study. In total, 2,255 FSW participated. The current study includes 2,220 participants whose age at initiation of selling sex for money was reported. Fifteen participants did not know, and nine refused to answer this question. Eleven participants who reported they were under age 18 when they started selling sex had inconsistent responses to other questions, including age at first sex. These individuals were therefore excluded from analyses.

### Data collection

Trained interviewers administered an in-person questionnaire and conducted rapid serological testing for HIV and syphilis based on national protocols, including pre- and post-test counselling. HIV testing was conducted using dual rapid testing (Alere Determine HIV 1/2 [Alere, Waltham, Massachusetts] and OraQuick HIV 1/2 [OraSure, Bethlehem, Pennsylvania]). Syphilis testing was conducted using rapid venereal disease research laboratory (VDRL) (rapid plasma reagin carbon, RapidGen Inc., Korea] titer and Treponema pallidum hemagglutination assays (Syphilis Rapid Test, DiaSpot Diagnostic, Indonesia) for detection of syphilis antibodies and classification of primary, active and past infection. Confirmatory testing with fluorescent treponemal antibody-absorption was not available. Subsequently, false-positive VDRL tests could not be excluded.

### Dependent variable

Selling sex during adolescence was defined as first having sex for money before the participant was 18 years old. Though Cameroon’s age of civil majority is 21, this study uses age 18 as a cutoff based on international standards and for comparability with research in other settings.(A. B. Grosso, Shianne; Mothopeng, Tampose; Sweitzer, Stephanie; Nkonyana, John; Mpooa, Nkomile; Taruberekera, Noah; and Baral, Stefan., 2018; A. L. Grosso et al., 2015; Inguane et al., 2015; Loza et al., 2010; Parcesepe, L’Engle, et al., 2016)

### Potential factors associated with early initiation of selling sex

The participant’s current age was collected as a continuous variable. Education level was dichotomized to compare those who completed primary school or higher to those who did not.(Parcesepe, L’engle, et al., 2016) Participants were asked the city in which they lived (Yaoundé, Douala, Bertoua, Bamenda or Kribi). In the analyses, the reference group was participants from Yaoundé. Consistent condom use was defined as always using condoms with regular and casual clients. Recent experience of sexual violence was defined as reporting being forced to have unwanted sex in the previous six months. Participants were asked if they ever had a pap smear. Recent STI and HIV testing history were separately assessed as reporting a test in the past 12 months. Recent experience of STI symptoms in the past 12 months was assessed for each of the following symptoms: blisters or sores in the genital region or anus; pain or burning sensation during urination (dysuria); vaginal discharge or unusual white discharge (unusual amount, odor, color); vaginal bleeding outside of their menstrual period; genital warts; anal warts; abnormal mass or swelling around genital organs; or other symptoms. Participants who reported any of these symptoms were asked whether they were treated by a doctor or other healthcare provider.

### Analysis

The relationship between each variable (HIV and syphilis test results, age, education, city, condom use, sexual violence, pap smear, HIV and STI testing, and STI symptoms and treatment) and initiation of selling sex during adolescence was assessed with bivariate logistic regression analyses and logistic regression analyses controlling for current age. The analyses did not control for number of years selling sex because this variable was not significantly related to selling sex during adolescence (p=0.085). Both FSW who sold sex as adolescence and FSW who sold sex only as adults reported they had sold sex for about five years. Additionally, this survey question seemed to be poorly understood. Over ten percent of participants reported the number of years they sold sex as greater than their current age minus the age at which they started selling sex. Variables significantly associated with selling sex during adolescence in age-adjusted analyses were included in a multivariable logistic regression model. The Akaike information criterion (AIC) was used to select the most parsimonious multivariable model. Analyses were conducted using Stata 14 (College Station, Texas). Missing data were handled using listwise deletion. In the final regression model, the prevalence of missing data was 2.2%. RDS weighting was not used due to pooling data from multiple cities to obtain a sufficient sample size of FSW who started selling sex as adolescents.

## Results

### Descriptive

As shown in Table 1, participants’ mean current age was 30.1 (interquartile range: 23-36). Most completed primary school or higher (84.5%). Over three quarters always used condoms with clients (77.4%). Over ten percent experienced recent sexual violence (10.5%). Few ever had a pap smear (7.2%). Over half recently tested for HIV (59.0%). Less than one quarter recently tested for STIs (23.7%). Over half recently experienced STI symptoms (52.3%, 1,161/2,220). Of those with any recent symptoms, about half had recently tested for STIs (52.2%, 599/1,148, data not shown in table), and less than half (48.9%, 567/1,159) had their symptoms treated by a health provider. Syphilis and HIV prevalence were 8.3% (185/2,220) and 24.5% (542/2,213). Among those testing positive for syphilis, 44.9% (83/185) reported no STI symptoms, and among those with no STI symptoms 9.6% (102/185) tested positive for syphilis (data not shown in table). Selling sex during adolescence was reported by 11.5% (256/2,220) of FSW.

**Table 1.** Prevalence and correlates of selling sex as an adolescent among adult female sex worker study participants in five cities in Cameroon, 2015-2016^1^

|  | Sold sex <18 |  | Sold sex ≥18 |  | Total |  |
| --- | --- | --- | --- | --- | --- | --- |
|  | % | n/N | % | n/N | % | n/N |
| Mean age at the time of the study | 23.1 |  | 31.0 |  | 30.1 |  |
| Completed primary school or higher | 82.4% | 211/256 | 84.8% | 1665/1963 | 84.5% | 1876/2219 |
| City |  |  |  |  |  |  |
| *Yaoundé* | 28.9% | 74/256 | 24.9% | 489/1964 | 25.4% | 563/2220 |
| *Douala* | 12.5% | 32/256 | 21.6% | 424/1964 | 20.5% | 456/2220 |
| *Bertoua* | 21.1% | 54/256 | 12.2% | 239/1964 | 13.2% | 293/2220 |
| *Bamenda* | 11.3% | 29/256 | 15.8% | 310/1964 | 15.3% | 339/2220 |
| *Kribi* | 26.2% | 67/256 | 25.6% | 502/1964 | 25.6% | 569/2220 |
| Sold sex <18 |  |  |  |  | 11.5% | 256/2200 |
| Always used condoms with clients | 66.3% | 169/255 | 78.9% | 1543/1957 | 77.4% | 1712/2212 |
| Was forced to have sex in the past 6 months | 16.0% | 41/256 | 9.7% | 191/1962 | 10.5% | 232/2218 |
| Ever had a Pap smear | 3.5% | 9/256 | 7.6% | 149/1953 | 7.2% | 158/2209 |
| Tested for HIV in the past 12 months | 49.2% | 125/254 | 60.3% | 1180/1958 | 59.0% | 1305/2212 |
| Tested for STIs in the past 12 months | 15.9% | 40/252 | 24.8% | 479/1935 | 23.7% | 519/2187 |
| Past 12 month STI symptoms |  |  |  |  |  |  |
| *Blisters or sores in the genital region or in the anus* | 25.0% | 64/256 | 21.5% | 422/1964 | 21.9% | 486/2220 |
| *Pain or burning sensation when urinating* | 30.5% | 78/256 | 24.3% | 477/1964 | 25.0% | 555/2220 |
| *Vaginal discharge or unusual white discharge* | 34.0% | 87/256 | 30.2% | 593/1964 | 30.6% | 680/2220 |
| *Irregular bleeding from the vagina* | 7.4% | 19/256 | 6.4% | 125/1964 | 6.5% | 144/2220 |
| *Genital warts* | 10.9% | 28/256 | 8.0% | 158/1964 | 8.4% | 186/2220 |
| *Anal warts* | 0.8% | 2/256 | 1.8% | 35/1964 | 1.7% | 37/2220 |
| *Abnormal mass or swelling around the genital organs* | 1.2% | 3/256 | 3.5% | 69/1964 | 3.2% | 72/2220 |
| *Other* | 14.5% | 37/256 | 14.9% | 293/1964 | 14.9% | 330/2220 |
| *Any of these symptoms* | 53.9% | 138/256 | 52.1% | 1023/1964 | 52.3% | 1161/2220 |
| Symptoms treated by a health provider | 40.6% | 56/138 | 50.1% | 511/1024 | 48.9% | 567/1159 |
| Tested positive for syphilis in the study | 8.2% | 21/256 | 8.4% | 164/1964 | 8.3% | 185/2220 |
| Tested positive for HIV in the study | 15.0% | 38/254 | 25.7% | 504/1959 | 24.5% | 542/2213 |

<sup>1</sup> HIV=human immunodeficiency virus; STI=sexually transmitted infection

### Bivariate

The mean current age of participants who started selling sex as adolescents was younger than that of those who started selling sex as adults (23.1 v. 31.0 years). Selling sex during adolescence was less common among participants in Douala and Bamenda and more common among participants in Bertoua. Compared to FSW who started selling sex as adults, a lower proportion of FSW who started as adolescents reported consistent condom use with clients (66.3% v. 78.9%), ever having a pap smear (3.5% v. 7.6%), recent HIV testing (49.2% v. 60.3%), and recent STI testing (15.9% v. 24.8%). A higher percentage of women who started selling sex as adolescents experienced recent sexual violence (16.0% v. 9.7%) and recent dysuria (30.5% v. 24.3%). Among participants reporting any recent STI symptoms, a lower percentage of those who sold sex as adolescents were treated by a health provider (40.6% v. 50.1%). HIV prevalence was lower among those who sold sex as adolescents than among those who sold sex only as adults (15.0% v. 25.5%).

### Age-adjusted

As shown in Table 2, after adjusting for current age, education was negatively associated with initiation of selling sex during adolescence (age-adjusted odds ratio [aaOR]: 0.48, 95% confidence interval [CI]: 0.32-0.70). Recent blisters or sores (aaOR: 1.49, 95% CI: 1.08-2.08) and genital warts (aaOR: 1.99, 95% CI: 1.24-3.18) and testing positive for HIV (aaOR: 1.5, 95% CI: 1.05-2.40) were positively associated with initiation of selling sex during adolescence. Receiving treatment for STI symptoms and a pap smear were no longer associated with selling sex during adolescence.

**Table 2.** Logistic regression analysis of correlates of initiation of selling sex as an adolescent among adult female sex worker study participants in five cities in Cameroon, 2015-2016^2^

|  | Bivariate |  |  | Age-adjusted |  |  |
| --- | --- | --- | --- | --- | --- | --- |
|  | OR | 95% CI | P | aaOR | 95% CI | P |
| Mean age at the time of the study | **0.82** | **0.79, 0.84** | **<0.001** |  |  |  |
| Completed primary school or higher | 0.84 | 0.59, 1.18 | 0.319 | **0.48** | **0.32, 0.70** | **<0.001** |
| City |  |  |  |  |  |  |
| *Yaoundé* | Reference category |  |  |  |  |  |
| *Douala* | **0.50** | **0.32, 0.77** | **0.002** | **0.63** | **0.39, 1.00** | **0.048** |
| *Bertoua* | **1.49** | **1.02, 2.19** | **0.040** | 0.86 | 0.57, 1.30 | 0.486 |
| *Bamenda* | **0.62** | **0.39, 0.97** | **0.037** | 0.81 | 0.50, 1.31 | 0.391 |
| *Kribi* | 0.88 | 0.62, 1.26 | 0.486 | **0.63** | **0.43, 0.92** | **0.016** |
| Always used condoms with clients | **0.53** | **0.40, 0.70** | **<0.001** | **0.50** | **0.37, 0.68** | **<0.001** |
| Was forced to have sex in the past 6 months | **1.77** | **1.23, 2.55** | **0.002** | **1.68** | **1.13, 2.50** | **0.010** |
| Ever had a Pap smear | **0.44** | **0.22, 0.88** | **0.019** | 1.26 | 0.60, 2.66 | 0.540 |
| Tested for HIV in the past 12 months | **0.64** | **0.49, 0.83** | **0.001** | **0.64** | **0.48, 0.84** | **0.002** |
| Tested for STIs in the past 12 months | **0.57** | **0.40, 0.82** | **0.002** | **0.63** | **0.43, 0.91** | **0.013** |
| Past 12 month STI symptoms |  |  |  |  |  |  |
| *Blisters or sores in the genital region or in the anus* | 1.22 | 0.90, 1.65 | 0.202 | **1.49** | **1.08, 2.08** | **0.016** |
| *Pain or burning sensation when urinating* | **1.37** | **1.03, 1.82** | **0.032** | **1.68** | **1.23, 2.29** | **0.001** |
| *Vaginal discharge or unusual white discharge* | 1.19 | 0.90, 1.57 | 0.216 | 1.22 | 0.91, 1.64 | 0.184 |
| *Irregular bleeding from the vagina* | 1.18 | 0.71, 1.95 | 0.519 | 1.33 | 0.77, 2.27 | 0.305 |
| *Genital warts* | 1.40 | 0.92, 2.15 | 0.118 | **1.99** | **1.24, 3.18** | **0.004** |
| *Anal warts* | 0.43 | 0.10, 1.82 | 0.253 | 0.88 | 0.20, 4.00 | 0.874 |
| *Abnormal mass or swelling around the genital organs* | 0.33 | 0.10, 1.04 | 0.059 | 0.60 | 0.18, 2.00 | 0.409 |
| *Other* | 0.96 | 0.67, 1.39 | 0.844 | 0.99 | 0.67, 1.47 | 0.962 |
| *Any of these symptoms* | 1.08 | 0.83, 1.40 | 0.584 | 1.16 | 0.87, 1.53 | 0.308 |
| Symptoms treated by a health provider | **0.68** | **0.47, 0.98** | **0.038** | 0.89 | 0.61, 1.31 | 0.551 |
| Tested positive for syphilis in the study | 1.02 | 0.63, 1.64 | 0.936 | 0.72 | 0.43, 1.21 | 0.210 |
| Tested positive for HIV in the study | **0.51** | **0.35, 0.73** | **<0.001** | **1.59** | **1.05, 2.40** | **0.027** |

<sup>2</sup> HIV=human immunodeficiency virus; STI=sexually transmitted infection; CI=confidence interval; OR=odds ratio; aaOR=age-adjusted odds ratio

### Multivariable

In the multivariable analysis, as shown in Table 3, current age (aOR: 0.81, 95% CI: 0.78-0.84), education (aOR: 0.49, 95% CI: 0.33-0.74), condom use with clients (aOR: 0.58, 95% CI: 0.42-0.80), recent HIV testing (aOR: 0.71, 95% CI: 0.53-0.96), and recent STI testing (aOR: 0.65, 95% CI: 0.44-0.96) were negatively associated with initiation of selling sex during adolescence. Experiencing recent sexual violence (aOR: 1.74, 95% CI: 1.15-2.64), dysuria (aOR: 1.50, 95% CI: 1.08-2.10), and genital warts (aOR: 1.78, 95% CI: 1.08-2.94) were positively associated with initiation of selling sex during adolescence.

**Table 3.**
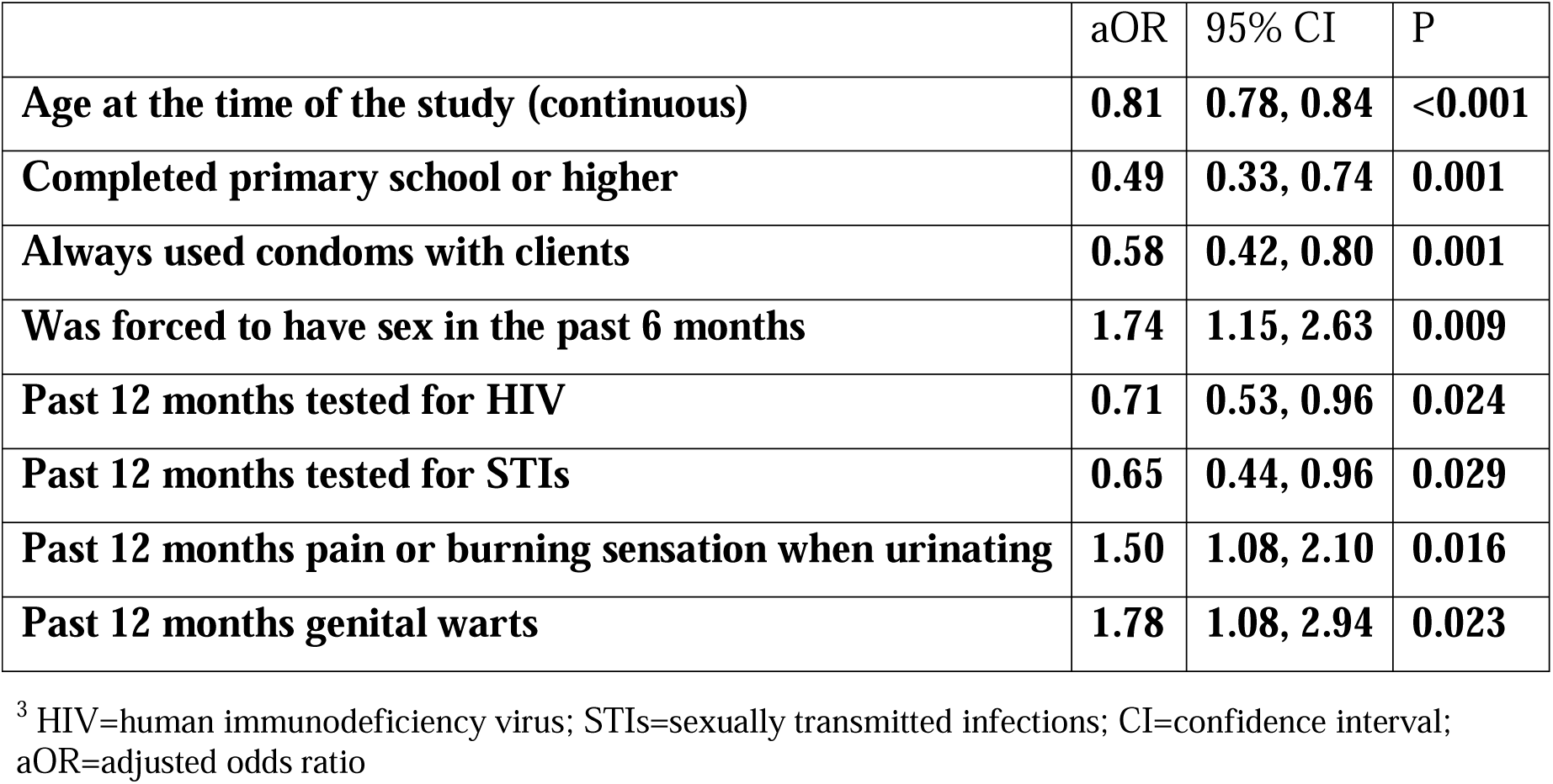
Multivariable logistic regression analysis of factors associated with selling sex as an adolescent among adult female sex worker study participants in five cities in Cameroon, 2015-2016^3^

## Discussion

Findings from this study give insight into differential HIV and STI-related acquisition and transmission risks among adolescent girls and young women. While some adolescents who sell sex likely stop and report other income sources as they get older, (Hounmenou, 2017b; Jonsson, Svedin, & Hydén, 2015) over one tenth of adult FSW in Cameroon continued selling sex after starting before the age of 18. Consistent with prior studies, even in age-adjusted and multivariable models adolescent initiation of selling sex was associated with heightened early and enduring vulnerability to STIs, as well as gaps in accessing testing and treatment services. As in other parts of the world,(Silverman, 2011) in Cameroon there is limited recognition that adolescents sell sex, resulting in little research with and no services for this population.(Mbassa Menick et al., 2009) Addressing adolescents in sex work is a critical harm reduction strategy. The development, implementation, and evaluation of evidence-based interventions to reduce risk for adolescents who sell sex and adult sex workers who started selling sex before the age of 18 is needed.

The symptoms of genital warts reported by FSW in this study may be an indication of human papillomavirus (HPV) infection, the most common cause of genital warts.(Garland, 2002) Cameroon has a high incidence of and mortality rate from cervical cancer, which is caused by HPV.(Bwaka & Dadjo, 2020) The prevalence of self-reported genital warts in this study was higher than prior studies using clinical diagnoses among FSW in Burkina Faso.(Low et al., 2011) The higher prevalence of genital warts among women who started selling sex as adolescents in this study underscores the importance of prevention, screening, and treatment of HPV and other STIs among this population.

The proportion of FSW in Cameroon who have received the HPV vaccination is unknown. In Cameroon HPV vaccinations were initially provided in schools, at clinics, and through mother-daughter approaches.(Ayissi et al., 2012; Ogembo et al., 2014) These approaches to vaccination may systematically miss some high-risk adolescents who sell sex given this study’s findings that FSW who sold sex as adolescents were unlikely to get tested for STIs and had lower odds of receiving any secondary school education. Adolescents who sell sex may also have been orphaned, left home, or been abused by their families, which would limit the feasibility of mother-daughter approaches.(A. L. Grosso, C; Diouf, D; Liestman, B; Ezouatchi, R; Thiam-Niangoin, M; Abo, K; Drame, FM; Bamba, A; Tety, J; Obodou, E; Ba, I; and Baral, S., 2017; Mbassa Menick et al., 2009) As HPV vaccination has been implemented more widely and at no cost to patients in Cameroon, misinformation has hindered uptake.(Bwaka & Dadjo, 2020) Vaccination coverage could potentially be improved through community engagement and outreach services.

Few FSW in this study had been tested for STIs or received a pap smear, particularly those who started selling sex as adolescents. In Cameroon syndromic management is more common than diagnostic testing for STIs as the result of the Ministry of Health’s national directive not to miss the opportunity from the first contact for presumptive treatment of a symptomatic STI to stop the transmission chain.(Nana Njamen T, 2017) This research contributes to better understanding the profile of asymptomatic patients. Improving access to adolescent- and key population-friendly STI diagnostic services could increase detection and treatment of asymptomatic STIs, such as those observed among some FSW in this study. Further research could assess whether implementing testing and treatment programs would be more cost-effective than relying on syndromic surveillance in this context.

Additionally, further research is needed to understand barriers to treating STIs among FSW who started selling sex as adolescents in Cameroon, such as stigma or costs.(Cook, 2020) Programs in Zimbabwe that have combined resources from interventions for adolescents and interventions for FSW to engage adolescents who sell sex in clinical services, health education and community mobilization could be considered for replication in Cameroon.(Busza et al., 2016)

Inconsistent use of condoms by clients and experience of sexual violence are STI risk mechanisms beyond the control of FSW, including those who sold sex as adolescents. In Benin, interventions targeting clients of sex workers increased condom use.(Lowndes et al., 2007) In South Africa increasing self-efficacy and condom negotiation power and educating FSW about violence prevention strategies decreased violence victimization and condomless sex.(Wechsberg, Luseno, Lam, Parry, & Morojele, 2006) In Ghana educating police about violence against FSW to reduce abuse and improve protection and access to justice has been implemented.(Asamoah, 2015) These interventions, if adapted for the Cameroonian context, could result in improved health outcomes among FSW and adolescents who sell sex. One promising program, Continuum of Prevention, Care and Treatment of HIV/AIDS with Most at-risk Populations in Cameroon, aims to reduce HIV and STIs and related morbidity and mortality in Cameroon through expanding gender-based violence screening and support services for FSW and other populations.

The focus of some funders on HIV testing yield over addressing STIs and prevention may exclude adolescents who sell sex or are at risk of selling sex because they tend to have lower HIV prevalence than older FSW who have been exposed to risk for a longer period of time.(Chikwari, Dringus, & Ferrand, 2018) Changing these priorities could improve engagement with adolescents and impact long-term sexual health outcomes.

This study’s findings should be considered in the light of several limitations. All data except HIV and syphilis test results were self-reported and may be affected by inaccurate recall or social desirability bias. Pain or burning sensation during urination is not specific to STI and could be a symptom of a non-sexually transmitted condition such as a urinary tract infection.(Ulmer, Gilbert, & De, 2014) To overcome limitations of assessing self-reported STI symptoms, further studies including clinical diagnostic testing for infections other than syphilis and HIV are warranted. These data were cross-sectional, which limits inferences about causality. However, entry into selling sex during adolescence occurred prior to many of the other variables assessed, as only adults participated in this study. The experiences of adult FSW who started selling sex as adolescents and continued likely differ from those who stopped selling sex before adulthood who would have been excluded from this study.(Mbassa Menick et al., 2009) Therefore further research, if legally and ethically feasible, working directly with adolescent girls who sell sex, including longitudinal studies following them into adulthood, are needed.

## Conclusions

Taken together, results from this study of sex workers in Cameroon demonstrate that entry into selling sex before the age of 18 is an indicator of longstanding risk for adverse sexual health outcomes.(Han, 2017) Prevention of adolescent girls selling sex could decrease the likelihood of forced sex, condomless sex, and STI acquisition. Among adolescent girls already selling sex, harm reduction measures should include HPV vaccination, gynecological examinations, STI testing, and treatment. These results reinforce the importance of empirically evaluating specific HIV and STI-related vulnerabilities among adolescent girls and young women to better ensure that programs address these specific risks to optimize potential impact.

## Data Availability

All data produced in the present study are available upon reasonable request to the authors.

## Acknowledgements

The authors thank the study participants for their time and for sharing their experiences and data to advance this research. We also acknowledge all study team members, advisors, and supporting staff from community-based organizations for their contributions to the study. Partners in Continuum of prevention, care, and treatment of HIV/AIDS with Most at-risk Populations project and those involved in the implementation of this study included CARE Cameroon, CARE USA, Johns Hopkins Bloomberg School of Public Health, Metabiota, Moto Action, the National AIDS Control Committee/Comité National de Lutte contre le Sida, Horizons Femmes, Humanity First, Alternatives, Alcondoms, Cameroon Medical Women’s Association, Cameroon National Association for Family Welfare, La Direction de la Recherche Operationnelle en Santé, L’Institut Nationale de Statistique, and L’Observatoire National de la Santé Publique du Cameroun. We also acknowledge the following collaborating health facilities: Yaoundé Military Hospital, Biyem-Assi District Hospital, Laquintinie Hospital, Douala Military Hospital, Nylon District Hospital, Centre Médical D’Arrondissement Soboum, Bertoua Regional Hospital, Bamenda Regional Hospital, and Kribi District Hospital. The authors thank the Cameroon government and in particular the Minister of Public Health, the Permanent Secretary of the National AIDS Control Committee, and their collaborators.

